# Clustered Phenotypes of Hypertensive Heart Disease With Strain Measurements Reveals Distinct Characteristics, Clinical Course, and Prognosis

**DOI:** 10.1101/2025.09.16.25335680

**Authors:** In-Chang Hwang, Hyue Mee Kim, Jiesuck Park, Hong-Mi Choi, Yeonyee E. Yoon, Goo-Yeong Cho

## Abstract

**Background:** Hypertensive heart disease (HHD) encompasses diverse clinical profiles, comorbidities, and cardiac remodeling, but current classifications insufficiently capture this heterogeneity or guide risk stratification.

**Methods:** We studied 1,607 patients with hypertension from the STRATS-HHD registry who underwent echocardiography at baseline and after 6–18 months of antihypertensive therapy. Twenty clinical, laboratory, and echocardiographic variables—including left atrial reservoir strain (LASr) and left ventricular global longitudinal strain (LV-GLS)—underwent principal component analysis and K-means clustering (K=4). Clusters were derived in a derivation cohort (n=1,204) and validated in an independent cohort (n=403). Longitudinal remodeling was assessed with baseline-adjusted models, and outcomes (CV death, heart failure hospitalization [HHF], coronary events, stroke, composite CV death/HHF, and major adverse cardiovascular events [MACE]) with multivariable Cox regression.

**Results:** Four clusters emerged: (1) AF-predominant, with advanced remodeling and highest event risk; (2) elderly, with metabolic–renal comorbidities but preserved function; (3) middle-aged, with prevalent coronary disease and relatively preserved function; and (4) younger, with severe hypertension, marked strain impairment, and greatest remodeling regression with therapy. Prognosis varied across clusters: cluster 1 had the highest risk of CV death, HHF, stroke, and MACE; cluster 3 had elevated coronary risk; and cluster 4 showed the most favorable outcomes. Medication–remodeling associations differed, with renin–angiotensin system blockade linked to LV mass regression in cluster 4.

**Conclusions:** Machine learning–based clustering incorporating LA and LV strain identified four distinct HHD phenotypes with divergent remodeling, therapeutic responses, and outcomes. Data-driven phenotyping may improve risk stratification and guide tailored management in hypertension.

## Introduction

Hypertension is the most common cardiovascular disease, leading to increased risk of mortality and morbidity.^1,2^ Its prevalence is rising, with recent studies estimating that one third to half of adults are affected or at risk.^1,3^ Although the hemodynamic consequences of hypertension are often summarized as increased afterload and vascular injury leading to target organ damage, the clinical profiles of patients are highly heterogeneous. This reflects diverse pathophysiological mechanisms—including arterial stiffness, salt sensitivity, sympathetic activation, and renin–angiotensin–aldosterone system dysregulation—as well as a wide spectrum of comorbidities such as diabetes, chronic kidney disease, dyslipidemia, coronary artery disease, arrhythmia, stroke, and heart failure.^4–6^ The chronicity of hypertension further complicates its course from early to advanced stages.^7–9^

Because of this diversity, management requires individualized approaches, which inevitably require classification of patients based on clinical context. Contemporary guidelines outline principles of antihypertensive therapy and provide specific recommendations for conditions such as diabetes, chronic kidney disease, or coronary disease.^3,10^ Yet in daily practice, therapeutic decisions often rely heavily on physicians’ judgment and experience rather than data-driven stratification.

Machine learning (ML) offers new opportunities to refine understanding of complex cardiovascular conditions. Unsupervised ML clustering has identified distinct phenotypes in valvular heart disease, heart failure, and cardiomyopathy.^11–13^ Several exploratory studies have applied clustering to hypertension and suggested its potential value for risk stratification and treatment guidance.^14–18^ However, patients with hypertensive heart disease (HHD) referred to tertiary centers—characterized by comorbidity burden and target-organ damage— remain understudied, despite their heightened vulnerability to cardiovascular complications.

In this study, we extended unsupervised ML clustering to a large tertiary-care cohort of patients with HHD who underwent echocardiography including automated left atrial (LA) and left ventricular (LV) strain measurements. By integrating strain with conventional echocardiographic parameters, we aimed to define comprehensive clinical clusters that capture the impact of hypertension on myocardial structure and function, and to evaluate their distinct characteristics, longitudinal course, and prognostic implications. We hypothesized that data-driven phenotyping would reveal clinically distinct subgroups of HHD with divergent remodeling trajectories and outcomes.

## Methods

### Study population

We included consecutive patients with hypertension who underwent baseline echocardiography at the time of diagnosis at Seoul National University Bundang Hospital (SNUBH) or Chung-Ang University Hospital (CAUH), two tertiary care centers in Korea, between 2006 and 2021. Eligible patients also had at least one follow-up echocardiogram obtained during antihypertensive therapy at 6–18-month intervals, as part of the Strain for Risk Assessment and Therapeutic Strategies in Patients With Hypertensive Heart Disease (STRATS-HHD) Registry.^19–21^ Exclusion criteria were: (1) specific cardiomyopathies, such as dilated cardiomyopathy, hypertrophic cardiomyopathy, restrictive cardiomyopathy, ischemic cardiomyopathy, stress-induced cardiomyopathy, Fabry disease, and MELAS (mitochondrial encephalopathy, lactic acidosis, and stroke-like episodes); (2) significant (≥moderate) valvular heart disease; (3) end-stage renal disease; (4) prior open-heart surgery, and (5) any cardiovascular diseases other than essential hypertension that could cause LVH (i.e., secondary hypertension).^20^ After applying these criteria, 1,872 patients were eligible. We then excluded patients with missing echocardiographic or laboratory data, including unavailable LA reservoir strain (LASr) or LV global longitudinal strain (LV-GLS), resulting in a final study population of 1,607 patients.

This study complied with the Declaration of Helsinki and was approved by institutional review boards of participating hospitals. Informed consent was waived owing to retrospective design and minimal risk. The study is registered at the Clinical Research Information Service of Korea (KCT0008091).

### Echocardiography

Comprehensive two-dimensional, M-mode, and Doppler echocardiography was performed using commercially available ultrasound systems with 2–2.5 MHz transducers. Measurements were obtained according to American Society of Echocardiography guidelines.^22^ LV end-diastolic and end-systolic dimensions, septal and posterior wall thicknesses were measured, and LV mass was calculated using the Devereux formula. LV mass index (LV-MI) was derived by normalizing LV mass to body surface area, with LV hypertrophy (LVH) defined as LVMI >115 g/m² in men or >95 g/m² in women.^23^ Relative wall thickness (RWT) was calculated as 2× posterior wall thickness ÷ LV end-diastolic diameter (LV-EDD); RWT >0.42 indicated concentric remodeling. LV ejection fraction (LV-EF) was calculated by Simpson’s biplane method from apical 2-and 4-chamber views. LA volume index (LAVI) was obtained by dividing LA volume by body surface area. Right ventricular systolic pressure was estimated from the peak velocity of tricuspid regurgitation plus an estimate of right atrial pressure.

### Automated measurement of LA and LV strain

For all included patients, LV-GLS and LASr were quantified using an artificial intelligence (AI)-based system (Sonix Health Workstation, Version 2.0; Ontact Health Co., Ltd., Korea) that provides fully automated view classification, segmentation, and parameter extraction.^24–27^ The algorithm automatically identified apical 4-, 2-, and 3-chamber views, as well as zoomed LV views when available. It then performed joint segmentation and motion estimation using a three-dimensional convolutional neural network trained in a semi-supervised framework, combining supervised learning for segmentation with unsupervised learning for motion dynamics. This process delineated LV endocardial and epicardial borders and LA endocardial borders in apical views. LV-GLS was calculated as the mean peak longitudinal strain from apical 4-, 2-, and 3-chamber views, referenced to the QRS complex. LASr was measured in the apical 4-chamber view as the first positive deflection relative to the QRS. When multiple views or cycles were available, the algorithm selected the one with the largest LV or LA area. Technical details and validation have been reported previously.^24^

### Outcomes

The primary outcomes were: (1) a composite of cardiovascular (CV) death and hospitalization for heart failure (HHF), and (2) major adverse cardiovascular events (MACE), defined as a composite of CV death, HHF, coronary events (unstable angina, myocardial infarction, or coronary revascularization), and stroke (ischemic or hemorrhagic). Each individual component of these composite endpoints was also assessed. HHF was defined as an inpatient admission for worsening signs and symptoms of HF. Clinical outcomes were ascertained through review of hospital records, structured telephone interviews, and linkage to national mortality databases.^28^

### Unsupervised machine learning-based clustering

To identify distinct phenotypes of HHD, we applied unsupervised machine learning–based clustering. Analyses were performed in R version 4.3.3 (R Foundation for Statistical Computing, Vienna, Austria).

Candidate clustering variables were prespecified and included demographics (age, sex, body mass index [BMI]), baseline blood pressure, comorbidities (diabetes, dyslipidemia, chronic kidney disease, atrial fibrillation, coronary artery disease), laboratory values (hemoglobin, estimated glomerular filtration rate, total cholesterol), and echocardiographic indices (LV end-diastolic volume [LV-EDV], RWT, LVMI, LV-GLS, E/e′ ratio, tricuspid regurgitant velocity, LAVI, and LASr). All variables were converted to numeric values. Missing data were imputed by variable-wise mean substitution, and continuous variables were standardized as z-scores before clustering.

Principal component analysis (PCA) was performed to evaluate collinearity and explore overall data structure. K-means clustering was then applied to the scaled dataset, with the number of clusters set to four (K=4) a priori based on clinical interpretability and model stability. Each patient was assigned to one of four phenotypic clusters. To illustrate distinctive features of each cluster, radar plots were constructed using the top discriminating variables.

The clustering model was derived from the SNUBH cohort and subsequently applied to the independent CAUH cohort for external validation. Outcomes were evaluated in both cohorts, including CV death, HHF, the composite of CV death or HHF, and major adverse cardiovascular events (MACE). Prognostic analyses were conducted both from the time of baseline echocardiography and from follow-up echocardiography. Kaplan–Meier survival curves were generated for each cluster, with differences assessed using log-rank tests.

### Statistical analysis

Continuous variables are summarized as mean ± standard deviation and categorical variables as counts with percentages. Between-group differences were assessed using Student’s t-test or Kruskal–Wallis test for continuous variables and χ² tests for categorical variables. Survival outcomes were analyzed using Kaplan–Meier curves with log-rank tests. Cox proportional hazards models were fitted with cluster assignment forced into the model; additional covariates were selected using Akaike Information Criterion–guided stepwise procedures. Proportional hazards assumptions were verified, and model discrimination was assessed by Harrell’s C-index. Pairwise hazard ratios (HRs) between clusters were derived from model-based contrasts.

Echocardiographic changes (Δ = follow-up – baseline) were compared using linear models adjusted for baseline values and prespecified covariates (age, sex, BMI, blood pressure, comorbidities, and medication use). Adjusted mean differences and 95% confidence intervals were obtained using estimated marginal means, with pairwise cluster comparisons corrected by the Tukey method. Associations between antihypertensive medications and reverse remodeling were assessed using linear models with robust (HC3) standard errors, adjusting for baseline values, cluster, cohort, and available covariates. Multiplicity was controlled with the Benjamini–Hochberg method.

All analyses were conducted in R version 4.3.3 (R Foundation for Statistical Computing, Vienna, Austria). A two-sided P<0.05 was considered statistically significant.

## Results

### Clinical characteristics

Among 1,872 participants in the STRATS-HHD registry, 1,607 with both LA and LV strain measurements and complete covariate data were included. The final cohort was divided into a derivation set (n=1,204, SNUBH) and a validation set (n=403, CAUH). Baseline characteristics are summarized in **Table 1**. The mean age was slightly higher in the validation cohort compared with the derivation cohort (66.3±12.8 vs. 64.7±13.1 years, p=0.031). Approximately two-thirds of patients were male (60.5% vs. 62.3%, p=0.576). The prevalence of diabetes (27.7% vs. 33.0%, p=0.047), dyslipidemia (26.3% vs. 32.5%, p=0.020), and coronary artery disease (34.4% vs. 50.1%, p<0.001) was significantly higher in the validation set. Baseline blood pressures did not differ significantly between the two cohorts (systolic blood pressure [SBP]: 152.9±24.7 vs. 152.8±21.4 mmHg, p=0.937; diastolic blood pressure [DBP]: 89.9±18.9 vs. 89.0±15.4 mmHg, p=0.340).

**Table 1.** Baseline characteristics.

| Variables | Derivation set<br>(n=1,204) | Validation set<br>(n=403) | P |
| --- | --- | --- | --- |
| **Clinical Factors** |  |  |  |
| Age (years) | 64.7 ± 13.1 | 66.3 ± 12.8 | 0.031 |
| Male sex | 729 (60.5%) | 251 (62.3%) | 0.576 |
| Diabetes mellitus | 333 (27.7%) | 133 (33.0%) | 0.047 |
| Dyslipidemia | 317 (26.3%) | 131 (32.5%) | 0.020 |
| Chronic kidney disease | 88 (7.3%) | 19 (4.7%) | 0.090 |
| Coronary artery disease | 414 (34.4%) | 202 (50.1%) | <0.001 |
| Atrial fibrillation | 164 (13.6%) | 61 (15.1%) | 0.499 |
| Stroke | 155 (12.9%) | 55 (13.6%) | 0.769 |
| **Anthropometric Factors** |  |  |  |
| Body mass index (kg/m<sup>2</sup>) | 25.2 ± 3.6 | 25.0 ± 3.6 | 0.331 |
| SBP at baseline (mmHg) | 152.9 ± 24.7 | 152.8 ± 21.4 | 0.937 |
| DBP at baseline (mmHg) | 89.9 ± 18.9 | 89.0 ± 15.4 | 0.340 |
| Heart rate (bpm) | 75.6 ± 15.2 | 74.0 ± 13.4 | 0.076 |
| **Laboratory Findings** |  |  |  |
| Hemoglobin (g/dL) | 13.4 ± 2.1 | 13.3 ± 2.1 | 0.293 |
| Blood urea nitrogen (mg/dL) | 17.9 ± 8.1 | 18.3 ± 10.8 | 0.544 |
| Serum creatinine (mg/dL) | 1.0 ± 0.6 | 1.1 ± 0.7 | 0.015 |
| GFR (mL/min/1.73m<sup>2</sup>) | 81.5 ± 25.9 | 73.1 ± 25.0 | <0.001 |
| Total cholesterol (mg/dL) | 169.4 ± 42.1 | 171.3 ± 43.3 | 0.438 |
| Triglyceride (mg/dL) | 132.7 ± 91.3 | 127.4 ± 95.0 | 0.354 |
| HDL cholesterol (mg/dL) | 45.6 ± 13.5 | 47.2 ± 12.5 | 0.048 |
| LDL cholesterol (mg/dL) | 103.3 ± 35.5 | 96.9 ± 29.9 | 0.002 |
| **Baseline echocardiography** |  |  |  |
| LV-EDD (mm) | 49.3 ± 6.5 | 48.0 ± 6.2 | <0.001 |
| LV-ESD (mm) | 32.7 ± 8.1 | 31.6 ± 7.3 | 0.012 |
| LV-EDV (mL) | 94.1 ± 38.4 | 91.4 ± 35.0 | 0.189 |
| LV-ESV (mL) | 43.5 ± 31.0 | 41.0 ± 26.5 | 0.126 |
| LV-EF (%) | 56.6 ± 12.1 | 57.2 ± 11.3 | 0.351 |
| LV-MI (g/m<sup>2</sup>) | 67.0 ± 30.1 | 71.0 ± 32.3 | 0.024 |
| RWT | 0.421 ± 0.097 | 0.437 ± 0.102 | <0.001 |
| LAVI (mL/m<sup>2</sup>) | 41.2 ± 18.9 | 38.4 ± 16.1 | 0.004 |
| Mitral E/e' ratio | 12.8 ± 6.1 | 12.8 ± 7.8 | 0.987 |
| LASr (%) | 23.9 ± 9.2 | 25.8 ± 9.5 | 0.001 |
| LV-GLS (%) | 14.6 ± 3.7 | 14.5 ± 3.5 | 0.606 |
| TR Vmax (m/s) | 2.4 ± 0.5 | 2.4 ± 0.4 | 0.299 |
| RVSP (mmHg) | 30.3 ± 9.6 | 29.4 ± 8.6 | 0.087 |
| **LV geometry at baseline** |  |  | <0.001 |
| Normal | 424 (35.2%) | 124 (30.8%) |  |
| Concentric remodeling | 222 (18.4%) | 116 (28.8%) |  |
| Concentric hypertrophy | 275 (22.8%) | 101 (25.1%) |  |
| Eccentric hypertrophy | 283 (23.5%) | 62 (15.4%) |  |
| **Antihypertensive medication** |  |  |  |
| RAS blockers | 822 (68.3%) | 252 (62.5%) | 0.040 |
| Beta blockers | 432 (35.9%) | 151 (37.5%) | 0.607 |
| DHP-CCB | 567 (47.1%) | 171 (42.4%) | 0.117 |
| NDHP-CCB | 62 (5.1%) | 39 (9.7%) | 0.002 |
| Thiazide | 183 (15.2%) | 79 (19.6%) | 0.047 |
| MRA | 103 (8.6%) | 21 (5.2%) | 0.038 |
| **Clinical outcomes\*** |  |  |  |
| Follow-up duration (months) | 62.0 ± 39.1 | 107.5 ± 53.7 | <0.001 |
| Cardiovascular death | 40 (3.3%) | 27 (6.7%) | 0.005 |
| Coronary revascularization | 95 (7.9%) | 73 (18.1%) | <0.001 |
| Stroke | 79 (6.6%) | 32 (7.9%) | 0.406 |
| Hospitalization for heart failure (HHF) | 97 (8.1%) | 39 (9.7%) | 0.364 |
| Composite of CV death or HHF | 125 (10.4%) | 55 (13.6%) | 0.088 |
| MACE (CV death, coronary, stroke, HHF) | 302 (25.1%) | 137 (34.0%) | 0.001 |

Values are presented as mean ± SD or n (%). P values compare derivation and validation sets.
Major adverse cardiovascular events (MACE) were defined as a composite of cardiovascular death, coronary revascularization, stroke, or hospitalization for heart failure (HHF).
Abbreviations: AF, atrial fibrillation; BMI, body mass index; CAD, coronary artery disease; CKD, chronic kidney disease; CV, cardiovascular; DBP, diastolic blood pressure; DHP-CCB, dihydropyridine calcium channel blocker; EF, ejection fraction; ESD, end-systolic diameter; ESV, end-systolic volume; GFR, glomerular filtration rate; Hb, hemoglobin; HDL, high-density lipoprotein; HHF, hospitalization for heart failure; LA, left atrial; LASr, left atrial reservoir strain; LAVI, left atrial volume index; LDL, low-density lipoprotein; LV, left ventricular; LV-EDD, LV end-diastolic diameter; LV-EDV, LV end-diastolic volume; LV-EF, LV ejection fraction; LV-ESD, LV end-systolic diameter; LV-ESV, LV end-systolic volume; LV-GLS, LV global longitudinal strain; LV-MI, LV mass index; MACE, major adverse cardiovascular events; MRA, mineralocorticoid receptor antagonist; NDHP-CCB, non-dihydropyridine calcium channel blocker; RAS blockers, renin-angiotensin system blockers; RVSP, right ventricular systolic pressure; RWT, relative wall thickness; SBP, systolic blood pressure; TC, total cholesterol; TG, triglyceride; TR Vmax, tricuspid regurgitation maximal velocity.

### Clustering of the derivation set

In the derivation cohort, unsupervised machine-learning clustering was performed using clinical, laboratory, and echocardiographic variables, including LASr and LV-GLS. Principal component analysis was used for dimensionality exploration, and K-means clustering was applied to the scaled dataset. Four clusters (K=4) were prespecified based on model stability and clinical interpretability (**Figure 1**). The distribution of clinical and echocardiographic characteristics across clusters is summarized in **Table 2**.

**Figure 1.**
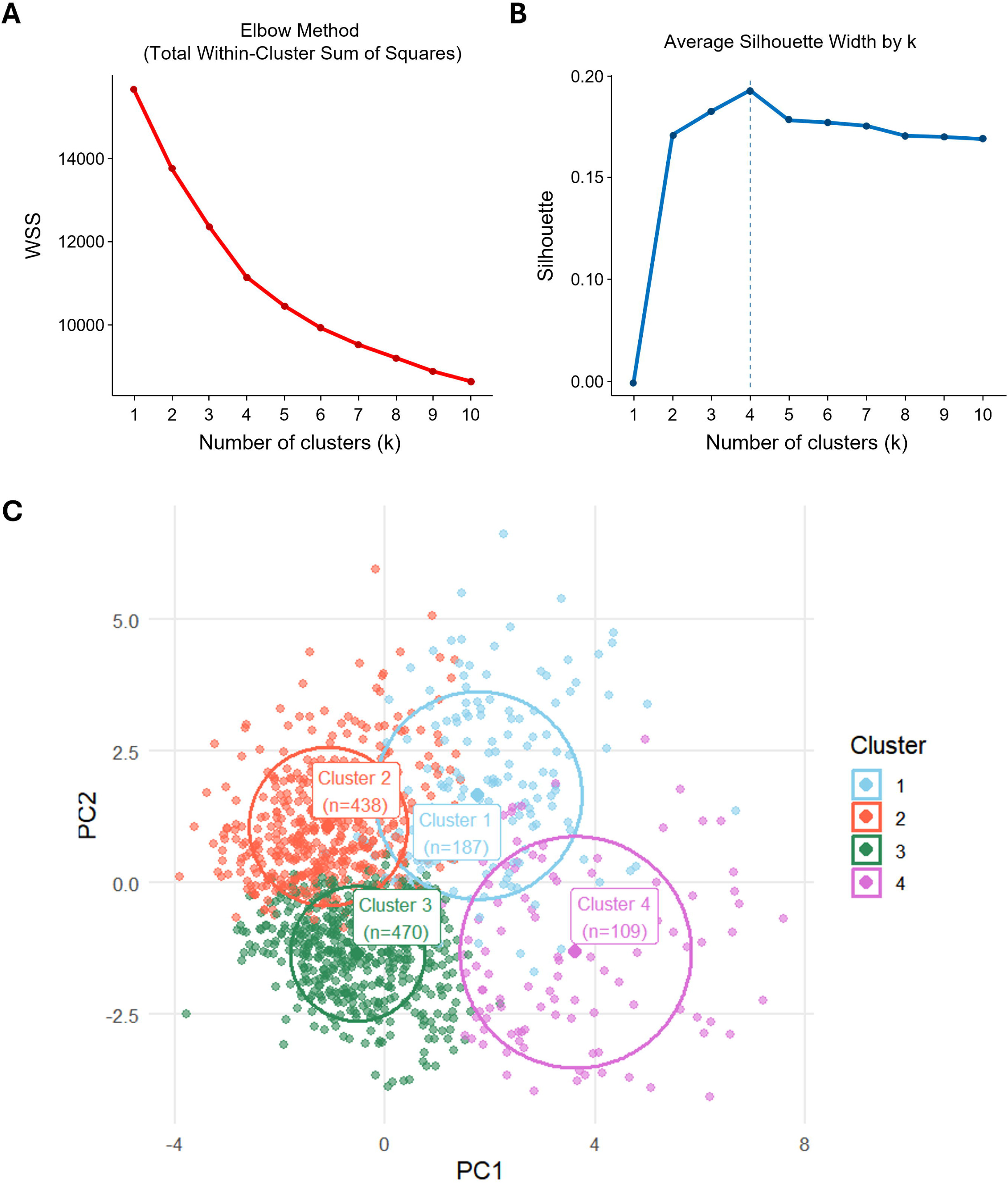
K-means clustering and PC1–PC2 scatter with cluster foci The optimal number of clusters was determined by **(A)** the elbow method (total within-cluster sum of squares) and **(B)** the silhouette method. **(C)** Two-dimensional scatter plots of individual patients, colored by cluster (cluster 1 = blue, cluster 2 = orange, cluster 3 = green, cluster 4 = purple), are shown. For each cluster, the filled circle indicates the centroid, and the surrounding ring depicts the root-mean-square (RMS) radius of dispersion. Axes show the first two principal components.

**Table 2.**
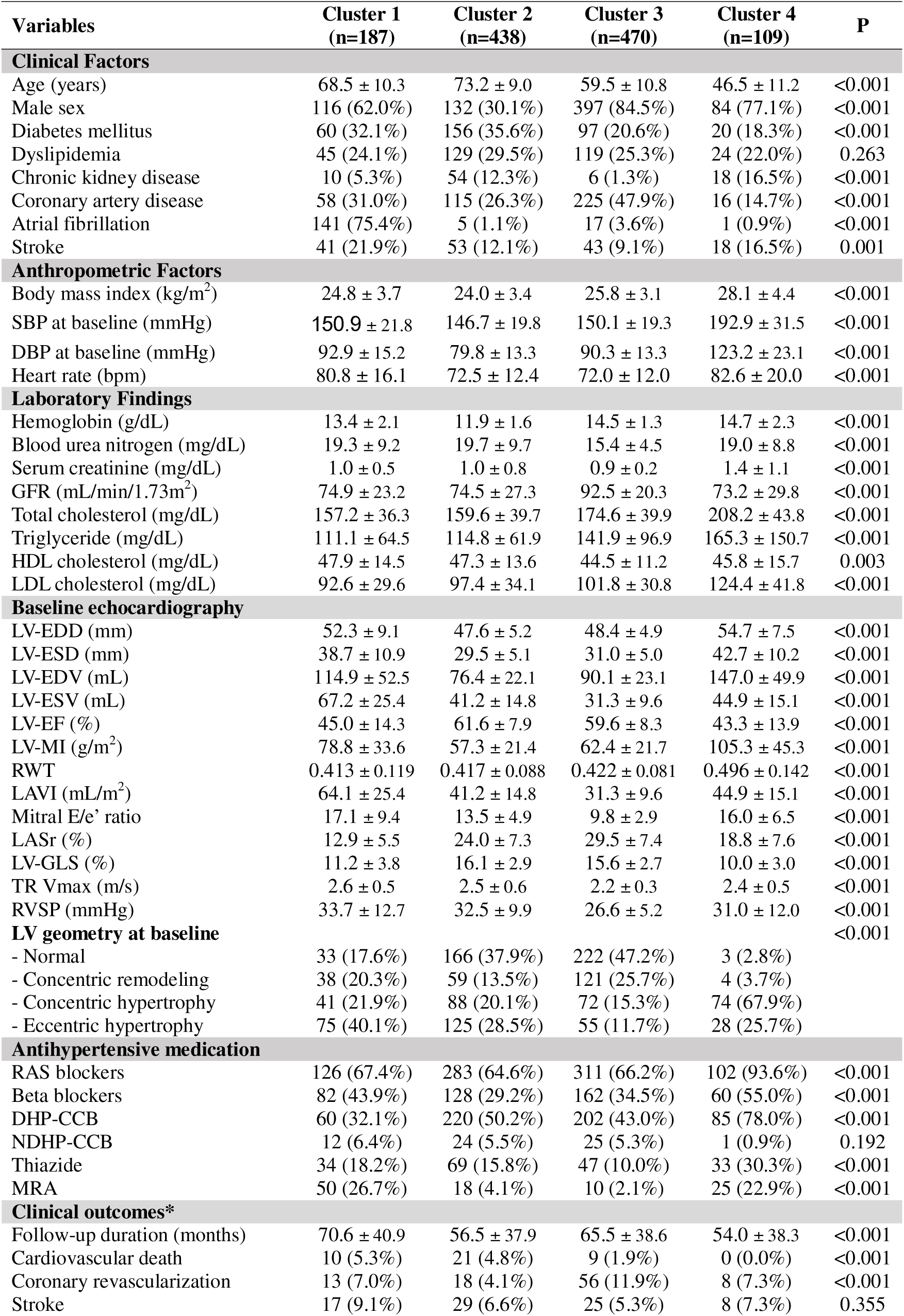
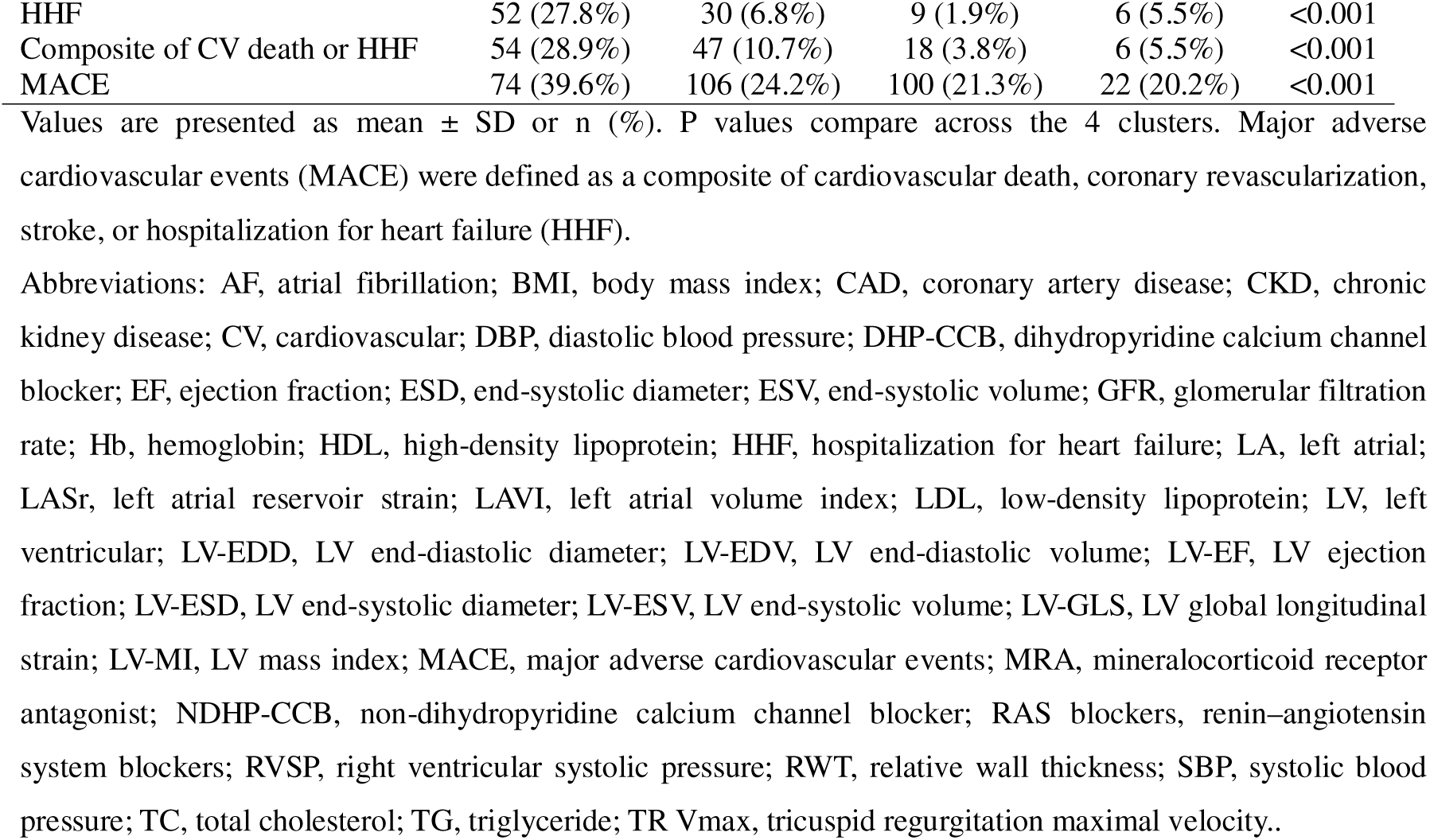
Clinical profile of clusters in derivation set.

Variable importance, assessed by K-means–consistent η² and random forest permutation analyses, identified seven major determinants of cluster assignment: history of atrial fibrillation, age, hemoglobin level, DBP, LASr, LV-GLS, and LAVI (**Supplementary Figure S2**). Radar plots constructed from these variables illustrated the distinct phenotypic signatures of each cluster (**Figure 2**).

**Figure 2.**
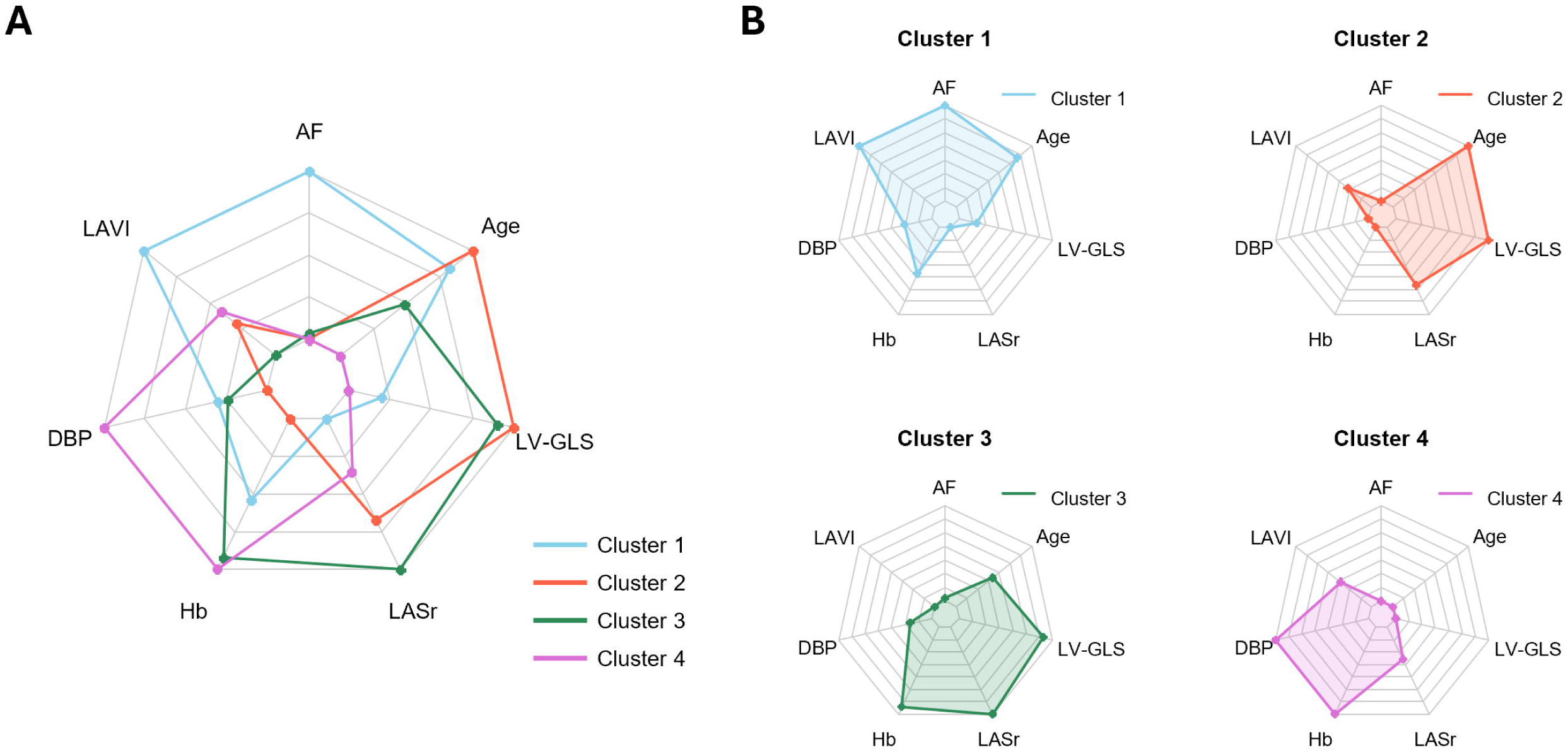
Radar plots of cluster phenotypes Radar plots summarize key clinical and echocardiographic features by cluster (e.g., age, diastolic blood pressure [DBP], hemoglobin [Hb], LA volume index [LAVI], LA reservoir strain [LASr], LV mass index [LVMI], LV global longitudinal strain [LV-GLS]). Axes are scaled comparably across panels, and line color denotes cluster assignment. LV-GLS is displayed as the absolute magnitude for interpretability.

Based on integrated clinical, laboratory, and echocardiographic data, as well as radar plot visualization, the four clusters exhibited distinct phenotypic signatures. Cluster 1 was characterized by a high prevalence of atrial fibrillation, marked LA enlargement, and pronounced reductions in both LASr and LV-GLS. Cluster 2 consisted of older individuals with lower DBP, smaller body size, impaired renal function, reduced hemoglobin levels, and preserved LV-GLS. Cluster 3 included middle-aged, predominantly male patients with a high prevalence of coronary artery disease and the most favorable functional profile, characterized by higher LASr and LV-GLS and smaller LAVI. Cluster 4 consisted of younger patients with elevated DBP and hemoglobin levels, the highest prevalence of LVH (particularly concentric LVH), moderately impaired LASr, mild but not severe LA enlargement, and markedly impaired LV-GLS.

### Changes in echocardiographic parameters according to the clusters

Across both cohorts, longitudinal changes in echocardiographic parameters after antihypertensive treatment (Δ = follow-up – baseline; median interval 10.2 months [IQR 6.8– 13.4]) were compared (**Figure 3**). In the derivation cohort, the most pronounced improvements were observed in cluster 4, which demonstrated significant reductions in LAVI, mitral E/e’ ratio, LV-EDV, and LV-MI, together with significant increases in LASr, LV-EF, and LV-GLS, compared to other clusters. Cluster 1 also exhibited modest improvements in LAVI, LASr, LV-EDV, LV-EF, and LV-GLS. Cluster 2 showed no significant interval changes in echocardiographic indices, while cluster 3 paradoxically demonstrated further increases in LAVI, TR Vmax, and LV-MI. Similar patterns were observed in the validation cohort.

**Figure 3.**
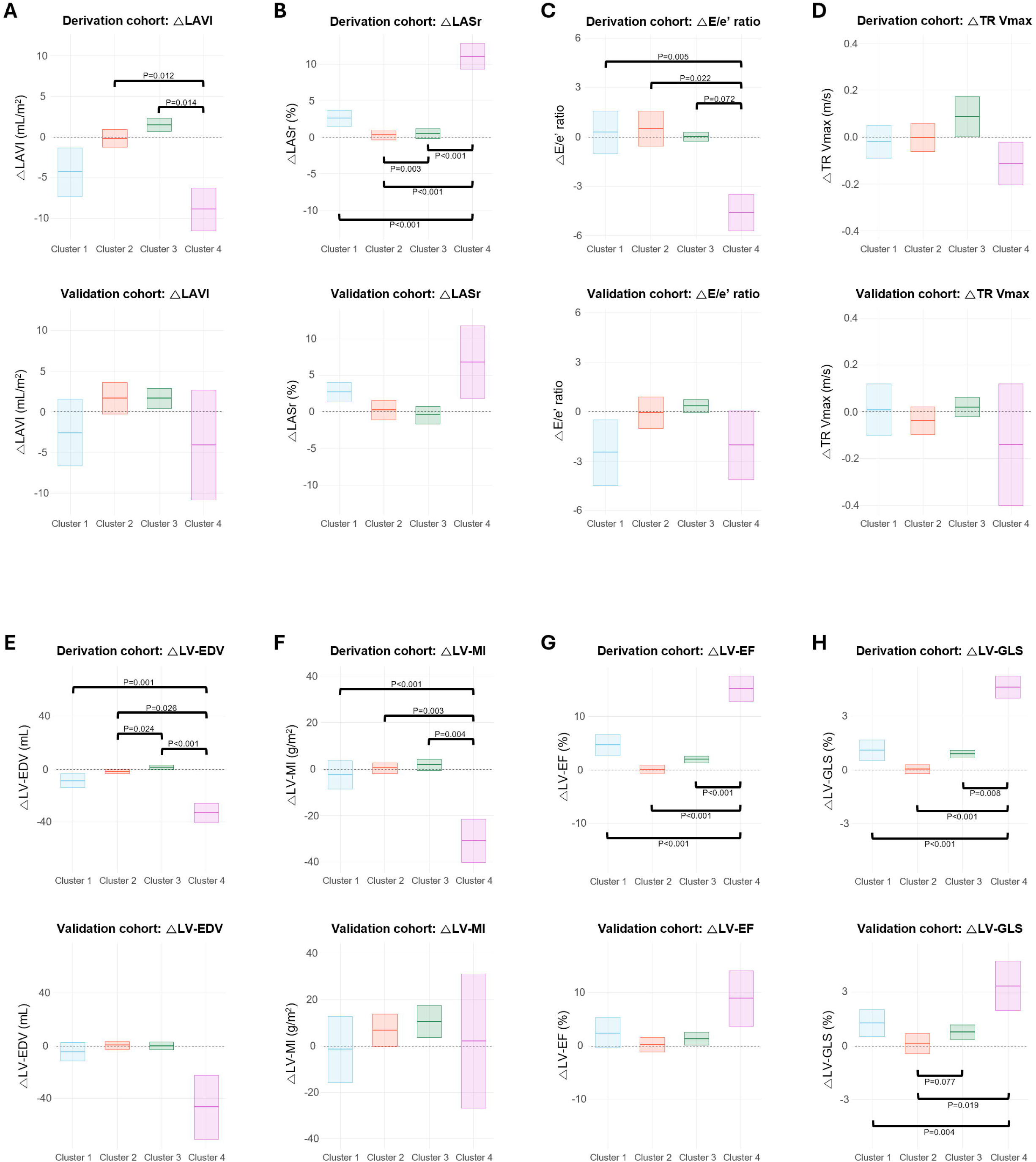
Changes in echocardiographic parameters by cluster Floating bars show mean Δ (follow-up − baseline) ± 95% confidence intervals for each cluster, stratified by cohort and parameter: **(A)** LAVI, **(B)** LASr, **(C)** E/e’ ratio, **(D)** tricuspid regurgitant velocity (TR Vmax], **(E)** LV-EDV, **(F)** LV-MI, **(G)** LV-EF, and **(H)** LV-GLS. Positive values indicate an increase at follow-up. LV-GLS was used as absolute values. Abbreviations: LAVI, LA volume index; LASr, LA reservoir strain; LV-EDV, LV end-diastolic volume; LV-ESV, LV end-systolic volume; LV-EF, LV ejection fraction; LV-MI, LV mass index; TR Vmax, tricuspid regurgitant velocity.

We further evaluated whether the class of antihypertensives influenced these remodeling trajectories (**Figure 4** and **Supplementary Figure S2**). Although most cluster-specific effects of drug classes were not prominent, several significant associations emerged. In cluster 1, calcium-channel blocker (CCB) use was associated with improved LV-EF, whereas mineralocorticoid receptor antagonist (MRA) use was linked to deterioration in LV-EF. In cluster 2, CCB use correlated with modest improvements in LASr and LV-GLS. In cluster 3, β-blocker therapy was associated with increased LAVI and reduced LASr. Notably, in cluster 4, thiazide use was associated with reduced LAVI and improved LASr, and renin– angiotensin system blockade (RASb) with LV-MI regression, whereas CCB therapy paradoxically correlated with increased LV-MI. Additionally, β-blocker and thiazide use in cluster 4 showed trends toward improved LV-EF and LV-GLS.

**Figure 4.**
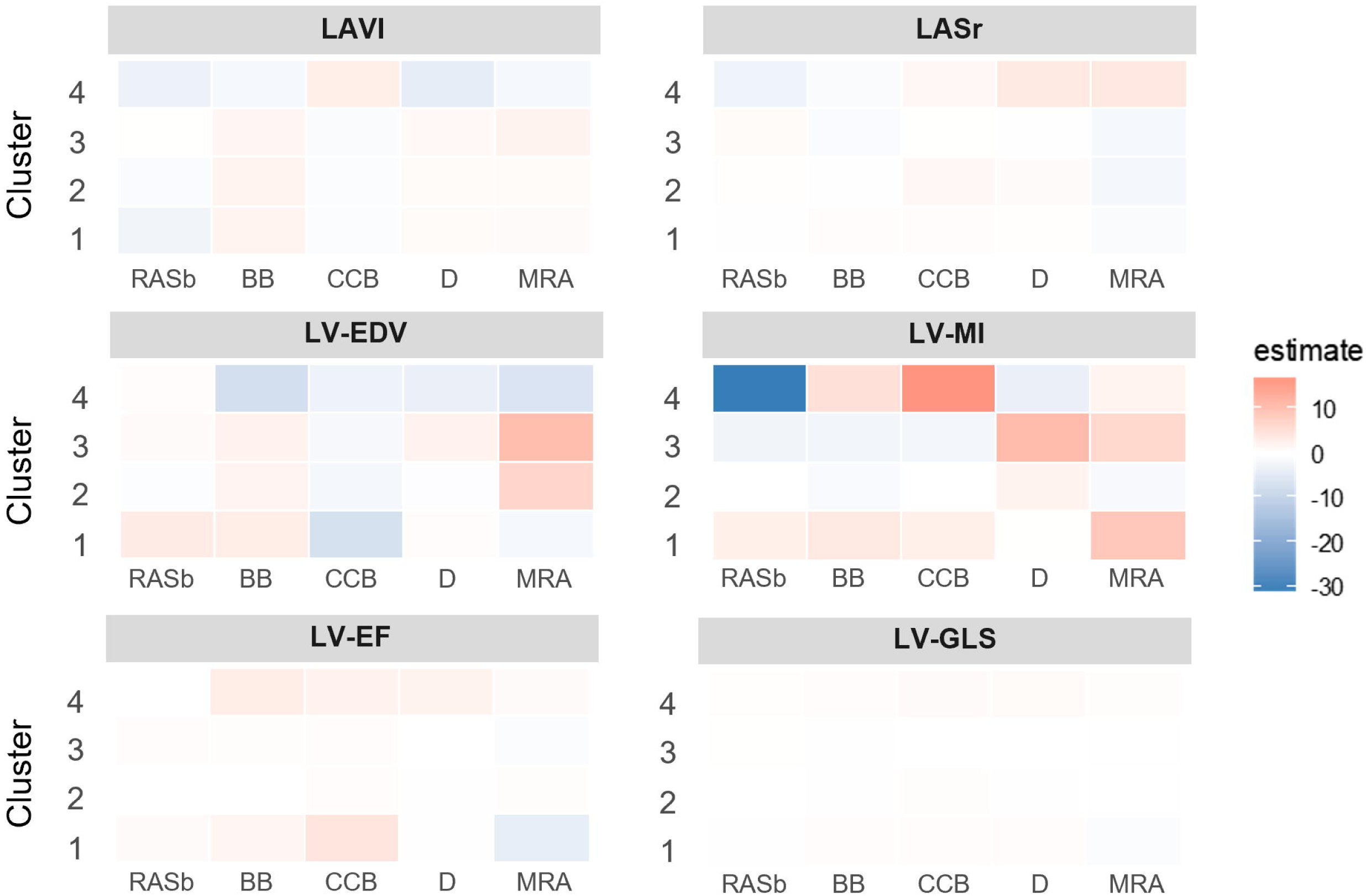
Medication effects on the changes in echocardiographic parameters by clusters Associations between antihypertensive medication classes (RAS blockers, β-blockers, calcium channel blockers [CCB], thiazide diuretics, and mineralocorticoid receptor antagonists [MRA]) and changes in echocardiographic parameters (LAVI, LASr, LVEDV, LVMI, LVEF, LV-GLS) are shown across clusters. The color scale indicates the direction of change (red = increase, blue = decrease), with transparency reflecting the magnitude of effect after adjustment. Abbreviations: LAVI, LA volume index; LASr, LA reservoir strain; LV-EDV, LV end-diastolic volume; LV-MI, LV mass index; LV-EF, LV ejection fraction; LV-GLS, LV global longitudinal strain; RAS, renin–angiotensin system; CCB, calcium channel blocker; MRA, mineralocorticoid receptor antagonist.

### Prognosis according to clusters

In order to assess the potential prognostic value of the ML-based phenotype clusters, the clinical outcomes, including CV death, coronary events, stroke, HHF, the composite of CV death or HHF, and MACE, were compared across the clusters in derivation and validation cohorts (**Figure 5** and **Table 3**). Cluster 1 experienced the highest risk of HHF and stroke, resulting in the greatest burden of the composite of CV death/HHF and overall MACE. Cluster 2 also showed increased CV death. By contrast, cluster 3 was characterized by a higher incidence of coronary events, while cluster 4 demonstrated the lowest risk of CV death and HHF. Overall, prognosis was poorest in cluster 1, intermediate in cluster 2, and most favorable in clusters 3 and 4.

**Figure 5.**
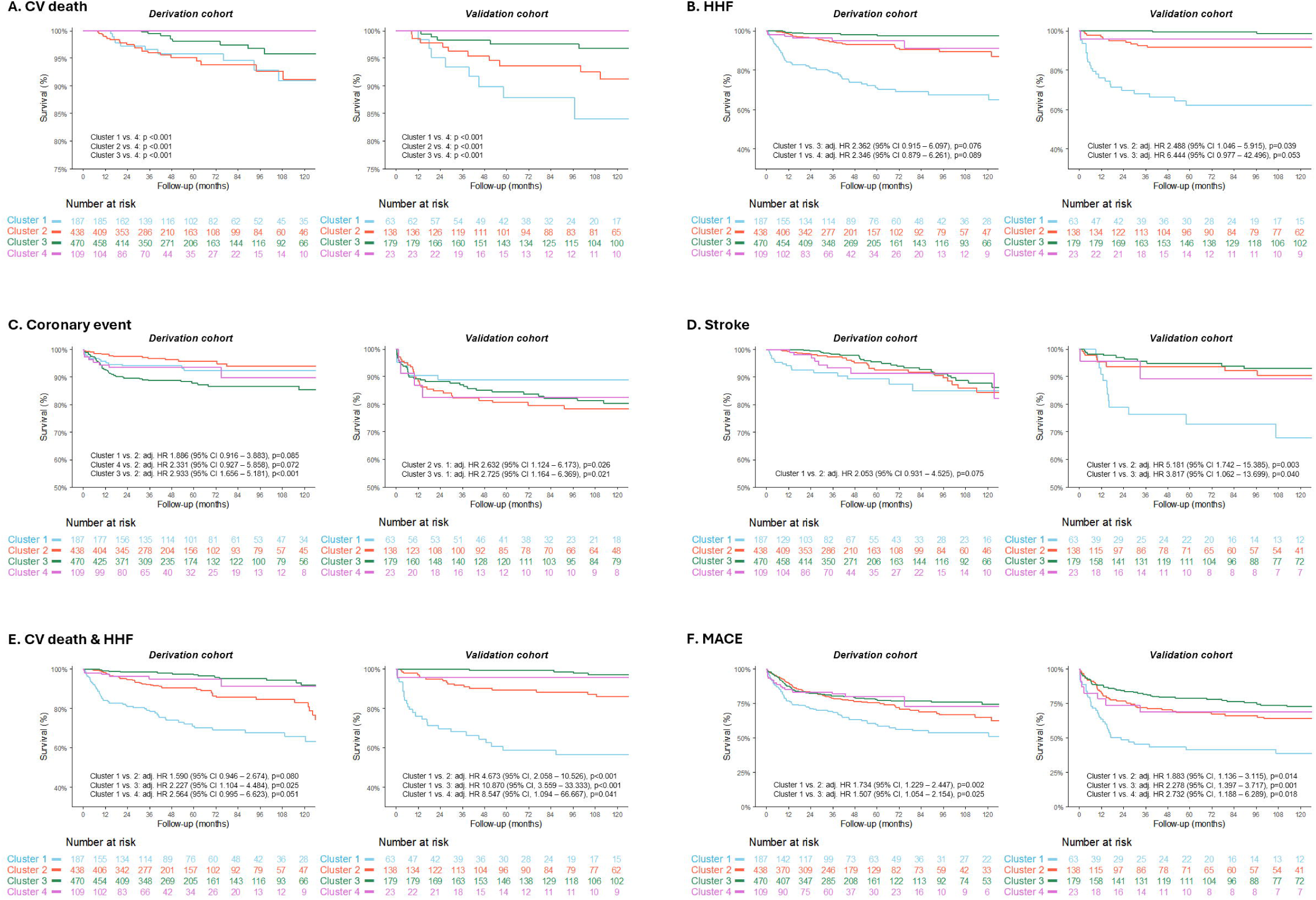
Survival analysis of clinical outcomes according to clusters Panels A–E show Kaplan–Meier curves for **(A)** CV death, **(B)** HHF, **(C)** coronary events, **(D)** stroke, **(E)** the composite of CV death or HF hospitalization, and **(F)** major adverse cardiovascular events (MACE). Curves are stratified by cluster (1 = blue, 2 = orange, 3 = green, 4 = purple). Adjusted HR, 95% CI, and p-values are shown for variables with p<0.100. Equivalent analyses are shown for the validation cohort to assess reproducibility of cluster-level risks. Abbreviations: CV, cardiovascular; HF, heart failure; MACE, major adverse cardiovascular events.

**Table 3.** Multivariable analyses for clinical outcomes.

|  | Derivation cohort |  | Validation cohort |  |
| --- | --- | --- | --- | --- |
|  | Adjusted HR (95% CI) | P-value | Adjusted HR (95% CI) | P-value |
| **CV death or HHF** |  |  |  |  |
| Age (per +1 year) | 1.044 (1.023 – 1.065) | <0.001 | 1.066 (1.033 – 1.099) | <0.001 |
| BMI (per +1 kg/m<sup>2</sup>) | 1.085 (1.033 – 1.140) | 0.001 | - | - |
| DM | 1.928 (1.339 – 2.776) | <0.001 | 2.024 (1.159 – 3.533) | 0.013 |
| CKD | 2.656 (1.595 – 4.422) | <0.001 | - | - |
| LASr (per +1%) | 0.962 (0.934 – 0.992) | 0.013 | 1.039 (0.999 – 1.081) | 0.056 |
| LV-GLS (per +1%) | 0.914 (0.865 – 0.966) | 0.001 | 0.911 (0.833 – 0.996) | 0.040 |
| Phenotype clusters |  |  |  |  |
| - Cluster 1 | *Referenced* |  | *Referenced* |  |
| - Cluster 2 | 0.629 (0.374 – 1.057) | 0.080 | 0.214 (0.095 – 0.486) | <0.001 |
| - Cluster 3 | 0.449 (0.223 – 0.906) | 0.025 | 0.092 (0.030 – 0.281) | <0.001 |
| - Cluster 4 | 0.390 (0.151 – 1.005) | 0.051 | 0.117 (0.015 – 0.914) | 0.041 |
| **MACE** |  |  |  |  |
| Age (per +1 year) | 1.029 (1.017 – 1.042) | <0.001 | - | - |
| SBP (per +1 mmHg) | 1.003 (0.998 – 1.008) | 0.289 | 1.008 (1.000 – 1.016) | 0.044 |
| BMI (per +1 kg/m<sup>2</sup>) | 1.052 (1.019 – 1.087) | 0.002 | 0.951 (0.904 – 1.001) | 0.054 |
| DM | 1.336 (1.046 – 1.706) | 0.020 | 2.161 (1.530 – 3.053) | <0.001 |
| TR Vmax (per +1 m/s) | - | - | - | - |
| LASr (per +1%) | 0.995 (0.978 – 1.013) | 0.594 | - | - |
| LV-GLS (per +1%) | 0.974 (0.938 – 1.012) | 0.181 | 0.911 (0.833 – 0.966) | 0.040 |
| Phenotype clusters |  |  |  |  |
| - Cluster 1 | *Referenced* |  | *Referenced* |  |
| - Cluster 2 | 0.577 (0.409 – 0.814) | 0.002 | 0.531 (0.321 – 0.880) | 0.014 |
| - Cluster 3 | 0.664 (0.464 – 0.949) | 0.025 | 0.439 (0.269 – 0.716) | 0.001 |
| - Cluster 4 | 0.728 (0.395 – 1.340) | 0.308 | 0.366 (0.159 – 0.842) | 0.018 |

Multivariable Cox proportional hazards regression was performed using an Akaike Information Criterion (AIC)-based stepwise selection method, with phenotype clusters forcibly included in all models. Hazard ratios (HRs) are shown with 95% confidence intervals (CIs).
Abbreviations: CV, cardiovascular; HHF, heart failure hospitalization; MACE, major adverse cardiovascular events; DM, diabetes mellitus; CKD, chronic kidney disease; LASr, left atrial reservoir strain; LV-GLS, left ventricular global longitudinal strain; SBP, systolic blood pressure; TR Vmax, tricuspid regurgitation maximum velocity.

## Discussion

In this study, we applied unsupervised ML-based clustering to a large cohort of patients with hypertension referred to tertiary hospitals who underwent echocardiography with LA and LV strain analysis at baseline and follow-up. Four distinct phenotypes of HHD emerged: (1) an AF-predominant cluster with advanced remodeling and impaired function, (2) an elderly cluster with metabolic–renal comorbidities and preserved LA and LV longitudinal function, (3) a middle-aged cluster with prevalent coronary disease and relatively favorable cardiac function, and (4) a younger cluster with severe hypertension, predominant concentric LVH, and marked impairment of LASr and LV-GLS. These clusters exhibited divergent remodeling trajectories under antihypertensive therapy: cluster 4 showed the most pronounced improvement, cluster 1 modest recovery, cluster 2 little change, and cluster 3 deterioration. Differences in treatment response by drug class were also noted, most prominently LV mass regression with RAS blockade in cluster 4. Prognostically, cluster 1 carried the highest risk of CV death, HHF, stroke and MACE; cluster 2 an intermediate risk; and cluster 3 a relatively higher burden of coronary events, whereas cluster 4 demonstrated more favorable outcomes. Collectively, these findings indicate that ML-based clustering can capture clinically meaningful heterogeneity in HHD, offering potential to guide treatment strategies and improve prognostic assessment in patients referred to tertiary care.

### Wide clinical spectrum of hypertension

Hypertension is one of the most prevalent chronic diseases worldwide, and most patients present with comorbidities such as diabetes, dyslipidemia, chronic kidney disease, or cardiovascular disease. Epidemiologic studies report prevalence rates of 15–30% for diabetes, 35–45% for dyslipidemia, 25–35% for chronic kidney disease, and about 20% for cardiovascular disease.^4–6^ These burdens are even greater in high-risk hypertension.^29^ Because of this frequent overlap, management often requires tailored approaches that balance the benefits and risks of different antihypertensive classes. In addition, the severity and chronicity of hypertension drive remodeling of cardiac structure and function—collectively termed HHD—which typically involves LA enlargement and LV concentric remodeling or hypertrophy, although eccentric patterns are also observed. Such changes are strongly linked to adverse outcomes and underscore the importance of strict BP control.

Despite this complexity, developing individualized treatment algorithms for every phenotype is impractical given the sheer prevalence and heterogeneity of hypertension. Current guidelines therefore recommend broadly applicable drug classes, with some preferential strategies for specific comorbidities.^3,10^ However, uncertainty persists regarding the optimal management of patients with advanced hypertension, multiple comorbidities, or overt HHD. In real-world practice, treatment decisions often rely on the attending physician’s discretion, guided by clinical reasoning and experience. In this regard, ML-based phenotype clustering may offer a feasible approach by systematically integrating diverse clinical information, explaining heterogeneity through data-driven reasoning, and facilitating tailored management.

### ML-based clustering phenotypes in hypertension

Unsupervised ML clustering not only defines overt phenotypic groups but also uncovers latent or under-recognized disease patterns not readily apparent to clinicians.^11,14–16,18^ By operating independently of human heuristics, ML-based clustering can identify novel associations between baseline phenotype and subsequent prognosis. Recent studies have applied this approach to hypertension. Katz et al. identified a subgroup of hypertensive patients with biomarker and imaging features consistent with the myocardial substrate for HFpEF, suggesting that clustering can reveal hidden risk of progression toward overt HF.^14^ In the SPRINT trial, Yang et al. demonstrated that distinct hypertensive phenogroups exhibited heterogeneous cardiovascular risk and variable benefit from intensive BP lowering.^15^ More recently, Rauseo et al. applied clustering to large-scale clinical and imaging data, showing that phenotypic subgroups stratify outcome risk beyond conventional classification.^17^ Vaura et al. uncovered a metabolically challenged hypertensive subgroup with excess cardiometabolic burden and adverse outcomes,^16^ while Oikonomou et al. used computational phenomaps across randomized trials to show that ML-derived profiles can identify patients most likely to benefit from intensive BP reduction.^18^ Together, these studies support clustering as a tool for risk refinement and potentially for tailoring therapy.

### Implications of phenotype clustering for high-risk patients with HHD

Our study extends this literature by focusing on high-risk hypertensive patients referred to tertiary institutions, a population in which no prior phenomapping evidence existed. Compared with earlier clustering studies, our analysis shares conceptual similarities—such as heterogeneous cardiovascular risk and divergent treatment responses—but also offers several important advances.

First, unlike prior work that relied primarily on clinical or metabolic variables,^14–18^ we incorporated advanced echocardiographic measures, namely LASr and LV-GLS, which provided sensitive markers of subclinical myocardial remodeling. This approach allowed us to capture phenotypes characterized not only by comorbidity burden but also by functional impairment of the heart. Second, whereas earlier studies largely focused on risk stratification, we evaluated longitudinal remodeling trajectories under antihypertensive therapy. We observed that cluster 4, despite severe baseline LVH and strain impairment, showed substantial improvement at follow-up, while cluster 3 paradoxically deteriorated, highlighting heterogeneity in remodeling responses. Third, we explored differential treatment associations across clusters. For instance, RAS blockade was linked to LV mass regression in cluster 4, whereas CCB therapy correlated with paradoxical LVMI increase in the same cluster but was associated with strain improvements in cluster 2. Although exploratory, these findings suggest that ML-defined phenotypes may inform drug selection in HHD, moving closer to precision medicine in hypertension.

Additionally, our findings provide new insights into prognosis across phenotypic subgroups. Cluster 1 carried the highest risk of HHF and stroke, likely reflecting its high prevalence of atrial fibrillation and severely impaired LA and LV strain. Cluster 2 showed intermediate outcomes, with excess CV death consistent with advanced age and the burden of metabolic–renal comorbidities. Cluster 3 had a relatively higher incidence of coronary events, explained by its high CAD prevalence and adverse lipid profile (high LDL cholesterol and the lowest HDL cholesterol). Cluster 4, despite the highest LDL cholesterol and severe LVH with impaired LV-GLS at baseline, consisted of younger patients who demonstrated marked improvement in echocardiographic parameters, ultimately experiencing the most favorable prognosis. Collectively, these findings indicate that ML-based clustering not only stratifies remodeling patterns and therapeutic responses but also delineates prognostic heterogeneity, thereby offering practical value for managing high-risk HHD.

### Role of LA and LV strain measurements for phenotypic assessment of HHD

Speckle-tracking strain imaging provides sensitive, quantitative measures of myocardial and atrial function that surpass conventional echocardiographic parameters in detecting subclinical dysfunction.^30–32^ LV-GLS and LASr directly capture the hemodynamic consequences of hypertension—namely, LV concentric or eccentric remodeling and LA enlargement with impaired reservoir function.^23^ By reflecting the functional integrity of myocardial and atrial tissue, these indices offer a more direct window into hypertensive remodeling than traditional measures such as LV-EF, wall thickness, or chamber dimensions.

In the present study, strain parameters emerged as key discriminators of cluster assignment beyond conventional parameters such as LV-EF, LV-EDV, and LAVI, and contributed to identifying phenotypes with markedly different remodeling responses and prognoses. Moreover, the use of automated AI-based strain analysis ensured reproducibility and scalability, supporting the feasibility of broader clinical application. The combination of automated strain echocardiography and ML-based unsupervised clustering may provide a paradigm shift in the assessment of hypertension and other cardiovascular diseases. Automated strain reduces the time and effort required for measurements, while ML-based clustering may capture risk stratification traditionally dependent on clinical expertise and logical reasoning. Together, they may facilitate application of optimized treatment strategies in a scalable fashion. Although further validation is needed in larger and more diverse cohorts, these findings illustrate the promise of combining automated imaging biomarkers with ML-driven clustering in clinical practice.

### Limitations

This study has several limitations. First, its retrospective observational design introduces the possibility of unmeasured confounding. Second, the study population consisted of patients referred to tertiary hospitals, which may lead to selection bias and limit generalizability to the broader hypertensive population. Third, the exploratory analyses of antihypertensive drug effects should be interpreted with caution, as medication choice was not prespecified but determined by treating physicians, resulting in imbalanced distribution across clusters and incomplete information on drug doses or adherence. Fourth, the timing of follow-up echocardiography was not standardized but varied among patients; however, the median interval between baseline and follow-up studies was 10.2 months, with a minimum of 6 months. Finally, although external validation was performed across two institutions, further validation in larger and more diverse populations is required to confirm the robustness of these phenotypic clusters.

## Conclusions

Unsupervised ML-based clustering of patients with HHD using LA and LV strain measurements identified four clinically distinct phenotypes with divergent remodeling trajectories and prognoses. These clusters underscore the heterogeneity of HHD and suggest that data-driven phenotyping may support more personalized treatment strategies. Prospective studies are needed to validate these findings and determine whether phenotype-guided management can improve clinical outcomes.

## Clinical Perspectives

### Competency in Medical Knowledge

- Hypertensive heart disease encompasses heterogeneous structural and functional remodeling patterns that are not captured by conventional classifications.
- ML-based clustering with LA and LV strain imaging combined with ML-based clustering identifies clinically distinct subgroups with differing remodeling trajectories, treatment responses, and outcomes.

### Translational Outlook

- Data-driven phenotyping may enable tailored management of hypertension by linking specific phenotypes to prognosis and therapeutic responsiveness.
- Prospective, multicenter studies are needed to validate these clusters and determine whether phenotype-guided treatment strategies improve patient outcomes.

## Abbreviations

AI: artificial intelligence
BMI: body mass index
CI: confidence interval
CV: cardiovascular
HHD: hypertensive heart disease
HHF: hospitalization for heart failure
LASr: left atrial reservoir strain
LAVI: left atrial volume index
LVH: left ventricular hypertrophy
LV-MI: left ventricular mass index
LV-GLS: left ventricular global longitudinal strain
MACE: major adverse cardiovascular events
ML: machine learning
PCA: principal component analysis
RWT: relative wall thickness

## Data Availability

All data produced in the present study are available upon reasonable request to the corresponding authors.

## Acknowledgments

We thank Lia Ju, a registered diagnostic cardiac sonographer (RDCS), and Eun-Ju Choi, a research nurse, for their dedication and valuable support in the conduct of this study.

