## Supplementary figures for "Clustered Phenotypes of Hypertensive Heart Disease With Strain Measurements Reveals Distinct Characteristics, Clinical Course, and Prognosis"

**Short Title:** Phenotype clustering of HHD

In-Chang Hwang, MD^a,b*^, Hyue Mee Kim, MD^c*^, Jiesuck Park, MD^a^, Hong-Mi Choi, MD^a,b^, Yeonyee E. Yoon, MD, PhD^a,b^, Goo-Yeong Cho, MD, PhD^a,b^

^a^ Department of Cardiology, Cardiovascular Center, Seoul National University Bundang Hospital, Seongnam, Gyeonggi;

^b^ Department of Internal Medicine, Seoul National University College of Medicine, Seoul;

^c^ Division of Cardiology, Department of Internal Medicine, Chung-Ang University Hospital, Seoul, South Korea

^*^ These two authors equally contributed to this work as co-corresponding authors**.**

**Supplementary Figure S1. Variable importance analysis**

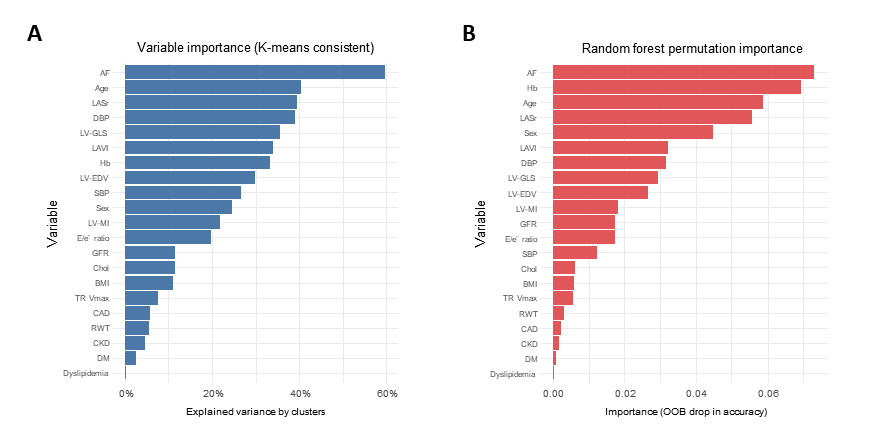

**(A)** K-means–consistent variable importance quantified by the proportion of between-cluster to total sum of squares (η² = BSS/TSS). **(B)** Random forest permutation importance, using out-of-bag (OOB) drop in accuracy as the metric, provided a complementary, model-based estimate of variable relevance.

**Supplementary Figure S2. Differential effects of antihypertensive medications on echocardiographic parameters across phenotype-clusters**

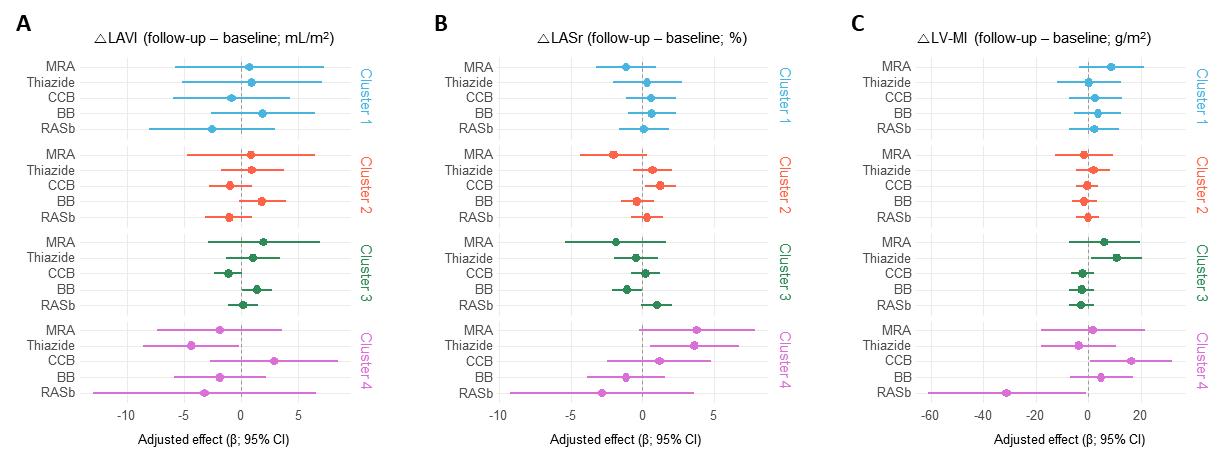

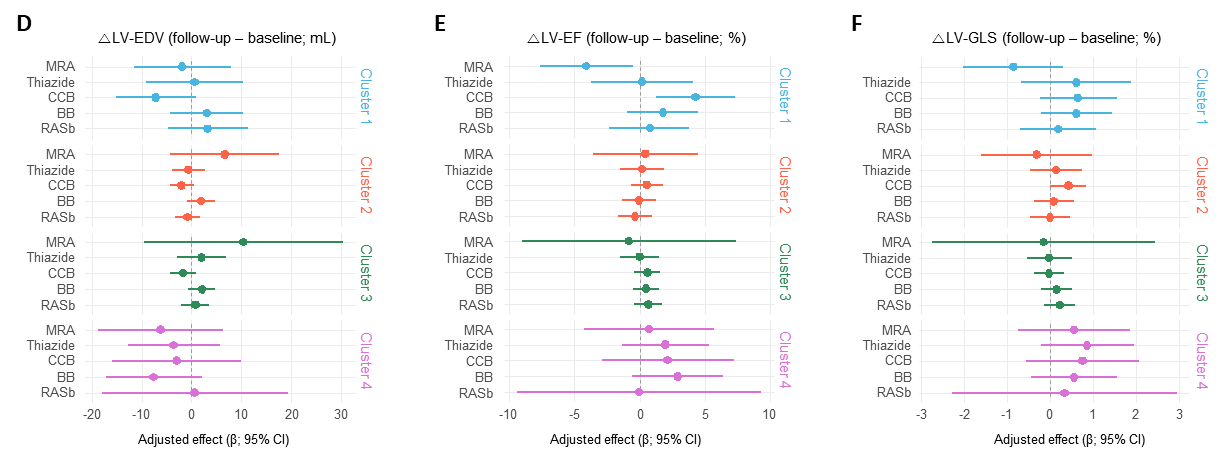

Forest plots illustrate the adjusted associations between antihypertensive medication use (renin–angiotensin system blockers, β-blockers, calcium-channel blockers, thiazide diuretics, and mineralocorticoid receptor antagonists) and longitudinal changes (Δ = follow-up – baseline) in echocardiographic indices of atrial and ventricular remodeling: **(A)** left atrial volume index (LAVI), **(B)** left atrial reservoir strain (LASr), **(C)** left ventricular mass index (LV-MI), **(D)** left ventricular end-diastolic volume (LV-EDV), **(E)** left ventricular ejection fraction (LV-EF), and **(F)** left ventricular global longitudinal strain (LV-GLS). Models were adjusted for baseline value, cohort, cluster assignment, and available covariates (age, sex, blood pressure, etc.), with robust (HC3) standard errors. Separate estimates are displayed across the four machine-learning–derived phenotype clusters.
